# Assessment of a bronchodilator response in preschoolers: a systematic review

**DOI:** 10.1101/2023.11.23.23298730

**Authors:** Matthew D. Wong, Kathleena Condon, Paul D. Robinson, Sadasivam Suresh, Syeda Farah Zahir, Peter D. Sly, Tamara L. Blake

## Abstract

**Background:** A bronchodilator response (BDR) can be assessed in preschool-aged children using spirometry, respiratory oscillometry, the interrupter technique, and specific airway resistance, yet a systematic comparison of BDR thresholds across studies has not been completed.

**Methods:** A systematic review was performed on all studies up to May 2023 measuring a bronchodilator effect in children 2-6 years old using one of these techniques (PROSPERO CRD42021264659). Studies were identified using MEDLINE, Cochrane, EMBASE, CINAHL via EBSCO, Web of Science databases, and reference lists of relevant manuscripts.

**Results:** Of 1224 screened studies, 43 were included. Over 85% were from predominantly Caucasian populations, and only 22 studies (51.2%) calculated a BDR cut-off based on a healthy control group. Sample sizes ranged from 25-916. Only two studies (4.6%) adhered to formal recommendations for study design: at least 300 subjects, randomised for placebo response testing in healthy children, and incorporated within-session and between-session test repeatability. A relative BDR was most consistently reported by the included studies (95%) but varied widely across all techniques. A variety of statistical methods were used to define a BDR. The highest BDR feasibility was reported with oscillometry techniques in this age group.

**Conclusion:** A BDR in 2-6-year-olds cannot be defined based on the reviewed literature due to inconsistent methodology and cut-off calculations. Precise and feasible evaluation of lung function in young children is crucial for early detection and intervention of airway diseases. A standardised approach is required to develop robust BDR thresholds.

## INTRODUCTION

Respiratory function testing enables the study of breathing mechanics and identifying airway and lung diseases. A positive bronchodilator response (BDR) is a key criterion used to diagnose and manage asthma, where a significant reversal of airflow limitation is demonstrated following acute treatment with a bronchodilator agent. However, there is no universally accepted definition of bronchodilator responsiveness in young children. Existing research on the diagnostic and prognostic value of BDR testing has produced conflicting results^1,2^.

One challenge with young children is the variable feasibility of available lung function techniques in ambulatory care^3^. Spirometry, which requires forced exhalation manoeuvres to achieve flow limitation, is often challenging for preschool-aged children to perform. Other techniques, such as oscillometry, specific airway resistance, and the interrupter technique, which can be performed during tidal breathing, offer greater feasibility for this age group but have varying clinical utility^4^. The clinical utility of bronchodilator testing depends on a low, within-session coefficient of repeatability (CR_intra_) between pre-bronchodilator (preBD) and post-bronchodilator (postBD) testing.

Before defining a significant change following bronchodilators, several characteristics of the pulmonary function test must be known^5^. First, the within-subject, within-session variation of the test, referred to as the coefficient of variability (CV), must be known for pre-bronchodilator and post-bronchodilator testing. Second, the within-subject CV for pre-placebo (prePL) and post-placebo (postPL) testing in healthy subjects provides information on the short-term repeatability of the technique. Lastly, the within-subject between-session coefficient of repeatability (CR_inter_) of BDR testing should be measured in age-matched subjects with and without disease. A clinically significant BDR should be greater than the within-session CV, greater than the test measurement error (i.e., beyond the 95% limits of agreement from placebo testing), and specific to the population (or condition) being tested^3,6^.

There are several ways to express a BDR. Reporting the absolute difference between preBD and postBD measurements (i.e., absolute BDR) favours taller children and may be influenced by large variations in preBD values^7,8^. An absolute BDR may be appropriate if the Bland-Altman mean-difference plot demonstrates that the measurement variability is independent of the mean^7^, as shown for spirometry in adults^9^. Reporting the relative BDR, calculated as a percentage change from the initial test (i.e., percentage of preBD), is better suited when the difference between postBD and preBD is proportional to the preBD value^10^. A relative BDR is less affected by the preBD values than an absolute BDR. If predictive values are available, reporting a BDR as a percentage of predicted or change in z-score eliminates the dependence on age, stature, and preBD values^6^.

Each lung function modality assesses different mechanical properties of the respiratory system. Spirometry assesses lung volume changes over time but depends on maintaining expiratory flow limitation until residual volume. The interrupter technique estimates alveolar pressure using a single-compartment model that assumes an equilibrium with mouth opening pressure. The specific airway resistance (sRaw) is estimated from tidal volume changes and gas flow. These three measures are based on a single-compartment model of respiratory mechanics to provide information about the resistive properties, predominantly airway resistance. Oscillometry uses linear dynamic system theory to model pressure, volume, and flow as functions over time during tidal breathing. Within oscillometry are several heterogeneous techniques that use pulses of square waves (impulse oscillometry), single-frequency sinusoidal waves (intra-breath oscillometry), or multiple frequencies of sinusoidal waves (spectral oscillometry).

Therefore, the definition of a BDR will vary depending on the lung function technique, disease process, and population being assessed. One method for defining a positive BDR is using the <5^th^ percentile or >95^th^ percentile of the response of healthy subjects to an inhaled bronchodilator^6,11^. Another approach is to account for the within-subject between-test repeatability in healthy subjects for the desired outcome variable where the BDR cut-off is calculated as the mean within-subject sample variance + (1.96 * within-subject sample standard deviation)^1^.

This systematic review explored the feasibility of BDR testing, the methods of BDR testing and reporting, and the derivation of BDR cut-off values across the following lung function techniques that have the most potential as safe, practical, and useful for testing in children 2-6 years old: spirometry, impulse oscillometry (IOS), intra-breath oscillometry (IB-OSC), spectral oscillometry (SPEC-OSC), interrupter technique, and specific airway resistance (sRaw).

## METHODS

### Search strategy

Following PRISMA guidelines, studies were identified from MEDLINE, Cochrane Central Register of Controlled Trials, EMBASE, the Cumulative Index to Nursing and Allied Health Literature (CINAHL) via EBSCO, Web of Science, and hand-searching reference lists of papers included in the review. The search text had bronchodilator response in children/paediatric population, using either spirometry, oscillometry (including forced oscillation technique, impulse oscillometry, within-breath oscillometry, and intra-breath oscillometry), interrupter technique, or specific airways resistance. The final searches from each database were performed in May 2023. Conference abstracts, reviews, editorials, and commentaries were not included due to the limited data available for assessment. Details of the entire search strategy are included in Table S1.

### Eligibility criteria

Included studies contained (1) a study population of children between 2-6 years old; (2) lung function involving spirometry, respiratory oscillometry, sRaw, or the interrupter technique; (3) a healthy control group; (4) use of a short-acting beta-2-agonist (SABA) administered by a metered dose inhaler and spacer; and (5) measurement of a bronchodilator response.

Studies were excluded if they included (1) a study population with no separate results provided for participants 2-6 years old; (2) a nebuliser-administered bronchodilator; (3) the bronchodilator was not a SABA; (4) or full-text manuscript was not available in English.

### Study selection and data extraction

Title and abstract screening, duplicate removal, full-text reviews, and data extraction from database searches were performed using Covidence systematic review software (Veritas Health Innovation, Melbourne, Australia). The authors (MDW, KC, and TB) reviewed the search references and identified potential inclusion studies based on titles, abstracts, and bibliographies. Three authors (MDW, KC, and TB) acquired and independently reviewed full-text studies meeting inclusion criteria. Two authors (MDW and KC) extracted data independently from all included studies. The data extracted from each study was the first author, publication year, study type, country, sample size (disease and control groups), population age, and lung function apparatus. Extraction of specific lung function outcomes preBD and postBD are outlined in Table S2. BDR data extracted included the feasibility, bronchodilator drug, bronchodilator dose, bronchodilator delivery method, the time between preBD and postBD testing, and whether pre-placebo (prePL) and post-placebo (postPL) testing was performed.

### Study quality assessment

The Joanna Briggs Institute (JBI) critical appraisal checklist for studies reporting prevalence data was used to assess studies included in the final review^12^.

### Data analyses

The feasibility of BDR testing was calculated by dividing the number of children with a successful BDR test by the number of children who attempted BDR testing and expressed as a percentage. No meta-analysis was planned due to the anticipated heterogeneity in BDR methodology and reporting for each lung function technique. Reported BDR cut-offs were summarised in terms of an absolute change (BDR_ABS_), a percentage of the initial test (BDR_%INIT_), a percentage of the predicted value (BDR_%PRED_), and z-score change (BDR_Z_).

## RESULTS

Our search conducted in May 2023 identified 2078 potential studies. After 854 duplicates were removed, 284 studies remained following title and abstract screening. The full-text review identified 242 studies for exclusion (Figure 1). Forty-three studies that met the pre-defined criteria were included for data extraction^1,8,13–53^.

**Figure 1.**
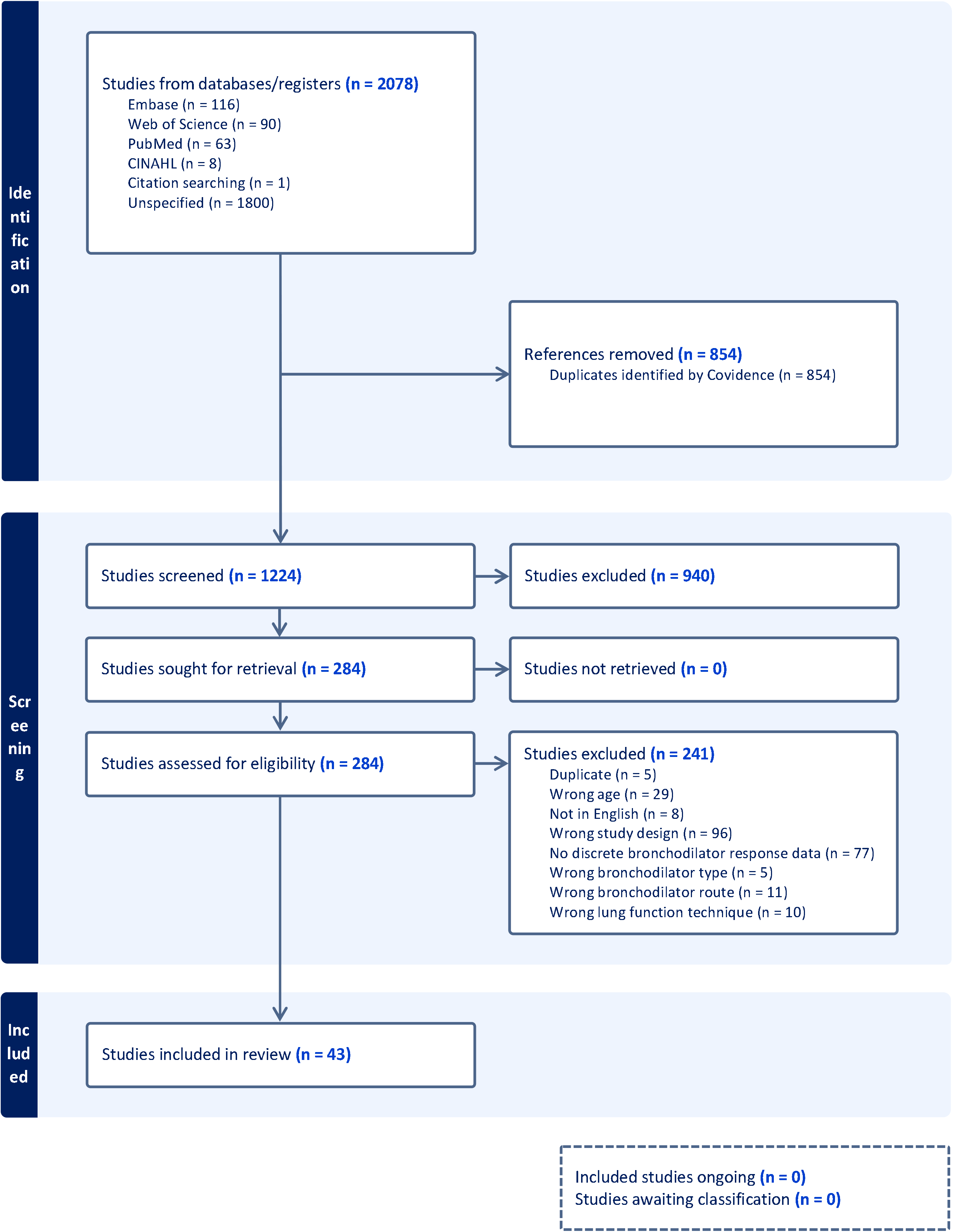
PRISMA diagram of study selection.

### Study characteristics

The demographics and descriptions of the 43 included studies are summarised in Table 1. Over 85% of the studies (n=37) were conducted in predominantly Caucasian populations. A large majority (81.4%) of the included studies consisted of cross-sectional study design, followed by cohort (9.3%) and case-control (9.3%). Of the included studies, 23 (53.5%) were published within the last ten years (2013-2023), and 20 (46.5%) were published >10 years ago. Thirty-one studies (72.1%) measured a BDR in healthy children (Table 1). Many studies utilised multiple lung function modalities, with fifteen studies (34.9%) using IOS, thirteen (30.2%) using spirometry, eleven (25.6%) using SPEC-OSC, ten (23.3%) using the interrupter technique, six (14%) using sRaw and one (2.3%) with IB-OSC. Details of the lung function equipment, bronchodilator testing methods and feasibility of bronchodilator response are summarised for each technique in the supplement (Tables S3-S7).

**Table 1.**
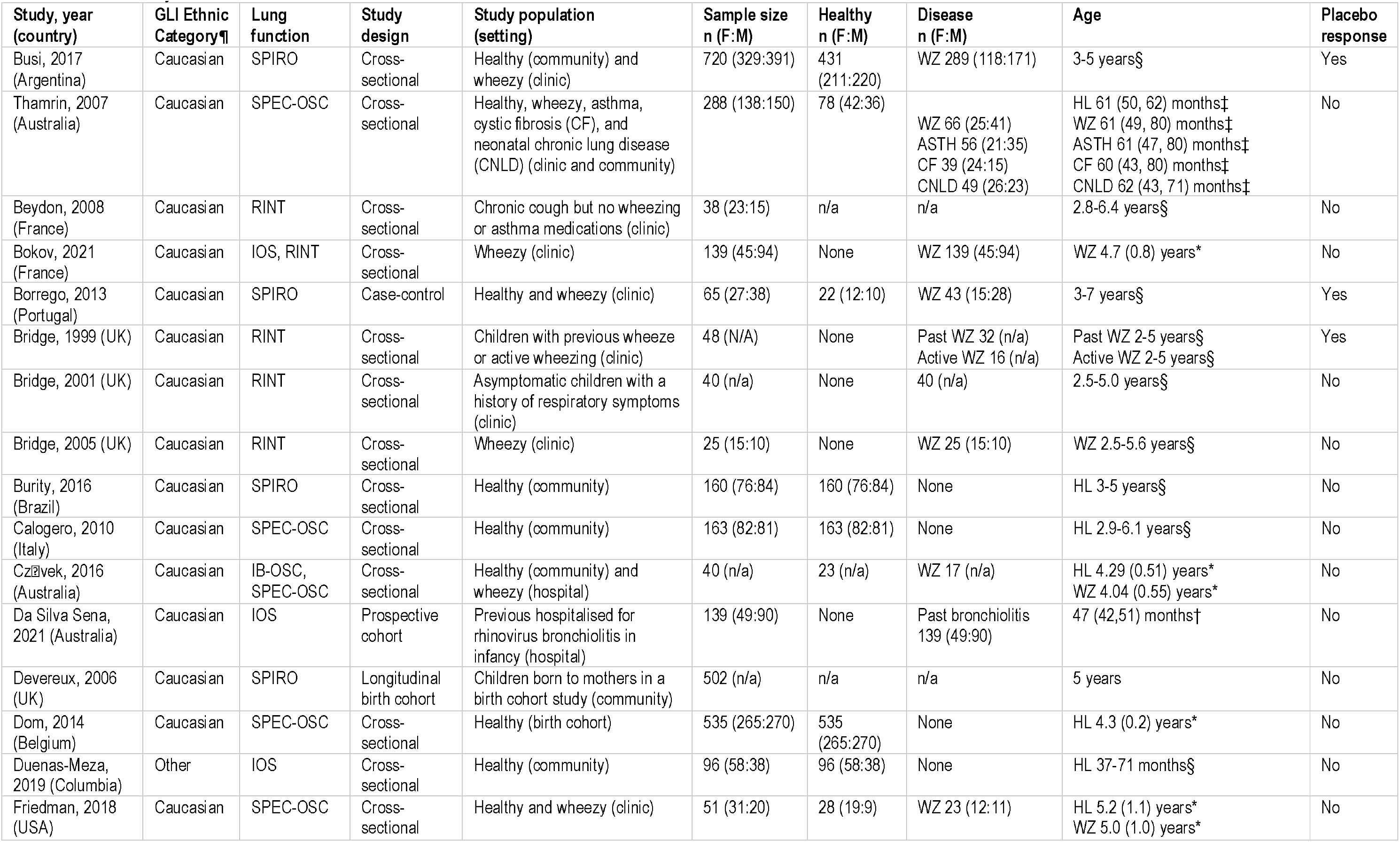
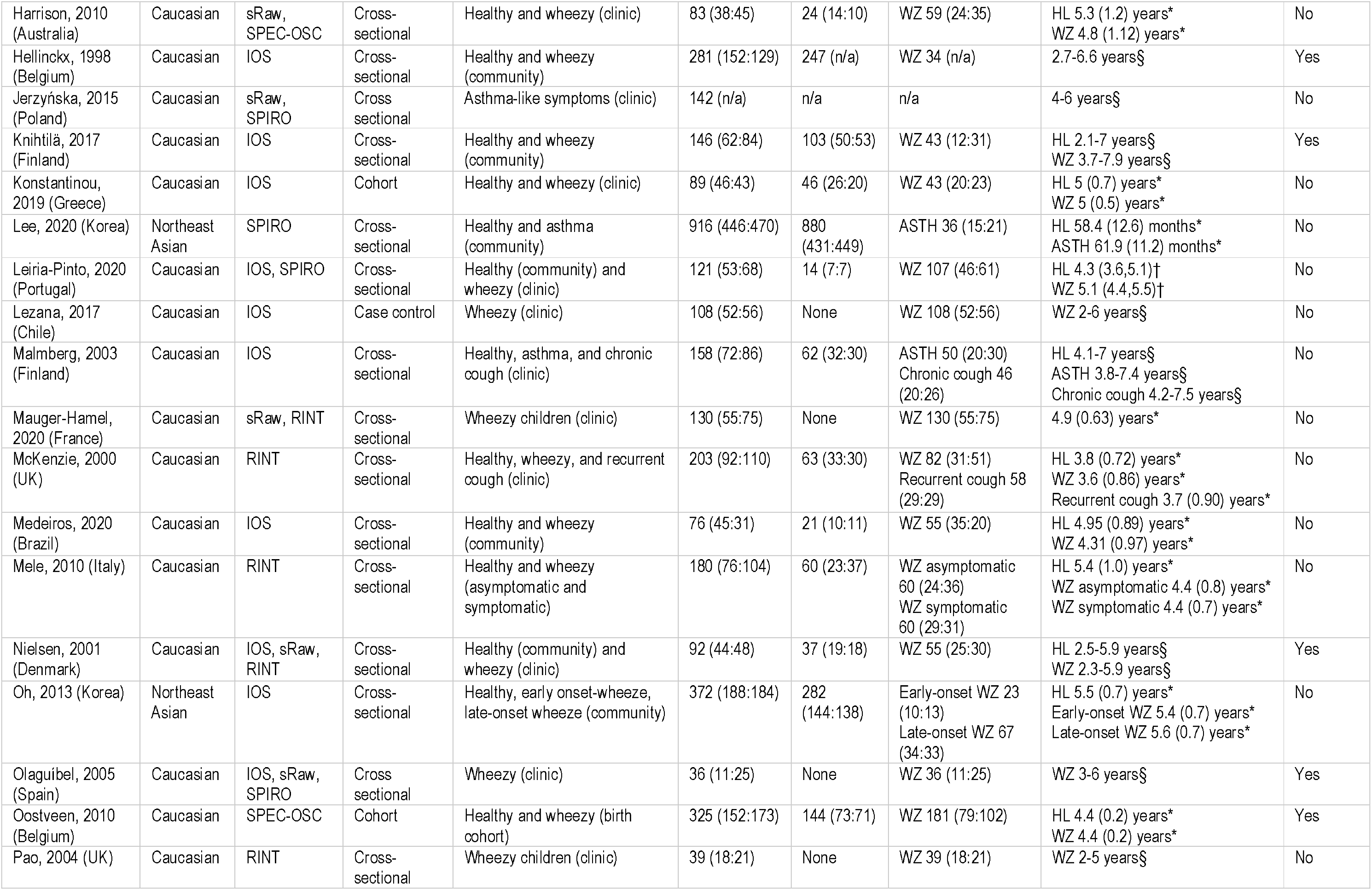
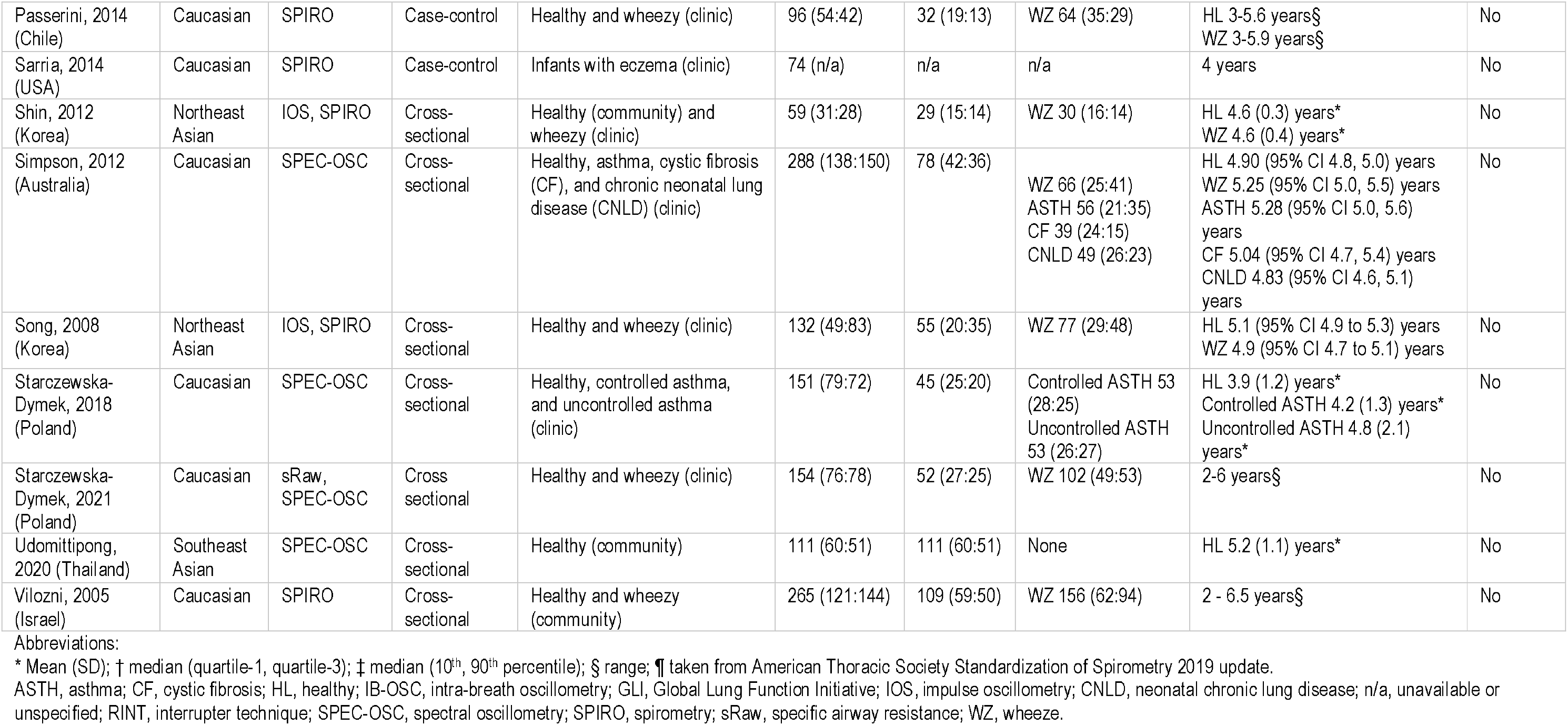
Summary of included studies.

### Quality Assessment

Twenty-nine studies (67.4%) included a healthy cohort, but only six (14%) had a healthy sample size of more than 300 children (Table S8). Only twenty-four studies (55.8%) proposed a BDR cut-off and included detailed methodology of BDR testing (e.g., bronchodilator dose, the time between preBD and postBD testing, and type of spacer used) (Table S8). Only nine studies (20.9%) controlled for the short-term repeatability of the lung function measurement with placebo response testing (Table S8). There were 24 studies (55.8%) explicitly stating how they derived a BDR cut-off from their sample.

### Feasibility of bronchodilator response testing by technique

The feasibility of bronchodilator response testing in preschool-aged children, including the bronchodilator drug type, drug dosage, sample size, and time between preBD and postBD measurements, is summarised in the supplementary tables. The highest median feasibility was reported for IOS and SPEC-OSC and the lowest for spirometry (Figure 2). In direct comparison within the same study, SPEC-OSC was more feasible in preschoolers (93%) than sRaw (68%)^27^.

**Figure 2.**
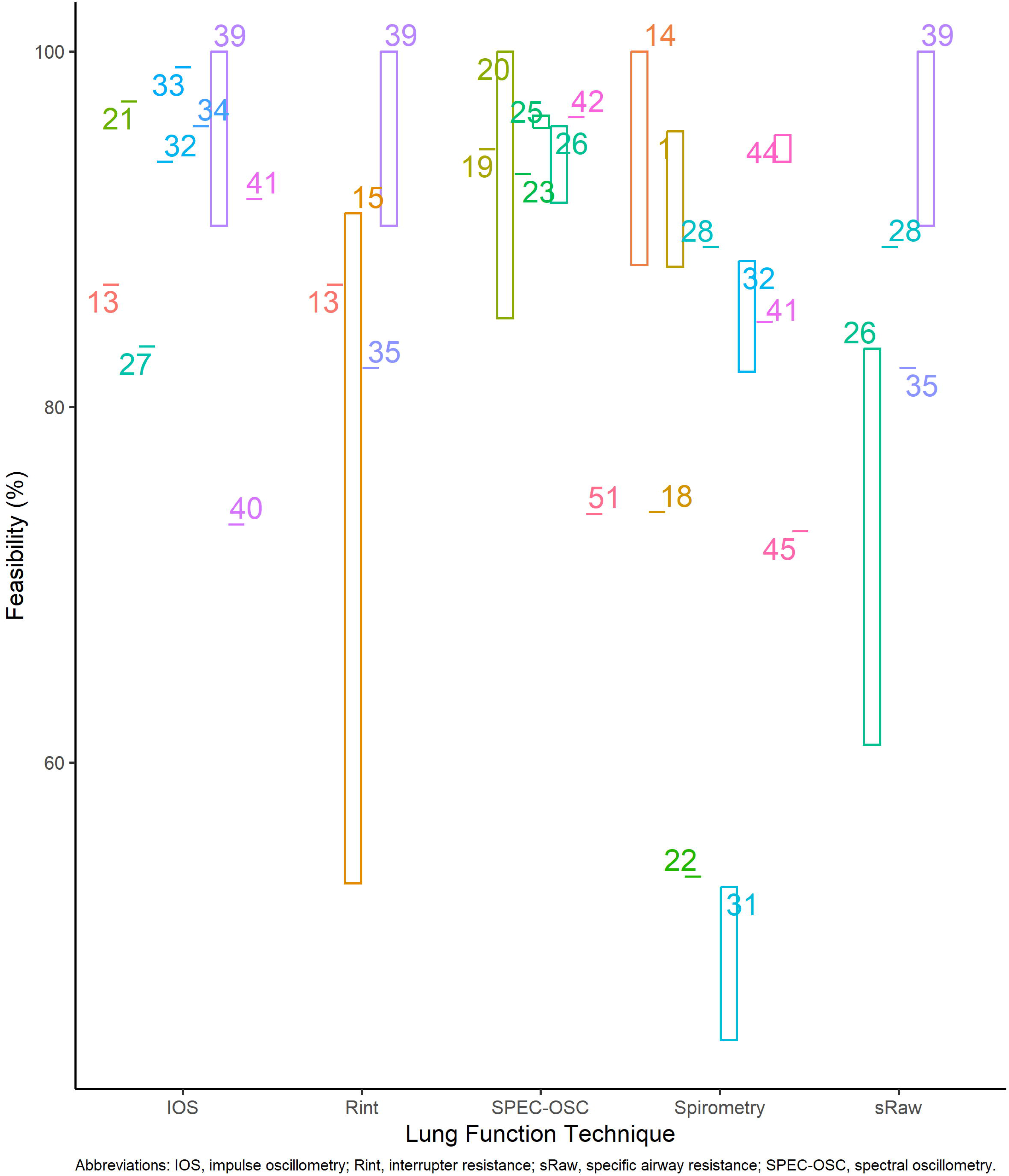
Reported feasibility for preschool bronchodilator response testing with colours representing individual referenced studies. The range of values is plotted in studies where feasibility is reported for subgroups within the overall cohort.

One study used terbutaline (2.3%), and the remainder used albuterol (11.6%) or salbutamol (86%). Doses of SABA varied between 200-600 mcg, and the time between preBD and postBD testing ranged between 10-20 minutes (Tables S3-S7).

### Defining a bronchodilator response

Twenty-two studies (51.2%) calculated a BDR cut-off based on a healthy control group (Table 2)^1,8,15,19,20,25,28,30–32,35,38–40,43,45,47–52^. BDR_%INIT_ was reported by all studies except Malmberg et al. 2003 for IOS outcomes and Nielsen et al. 2001 for sRaw (Figure 3). Four studies reported BDR_%PRED_^30,35,39,40^, and two reported BDR_Z_^30,39^. Only five studies deriving BDR cut-offs incorporated placebo testing of healthy subjects to account for the within-subject intrasession repeatability of the test^1,15,30,40,43^. The remaining 21 studies applied a BDR cut-off derived from published reference data to their study population. Results within individual techniques are discussed below.

**Table 2.**
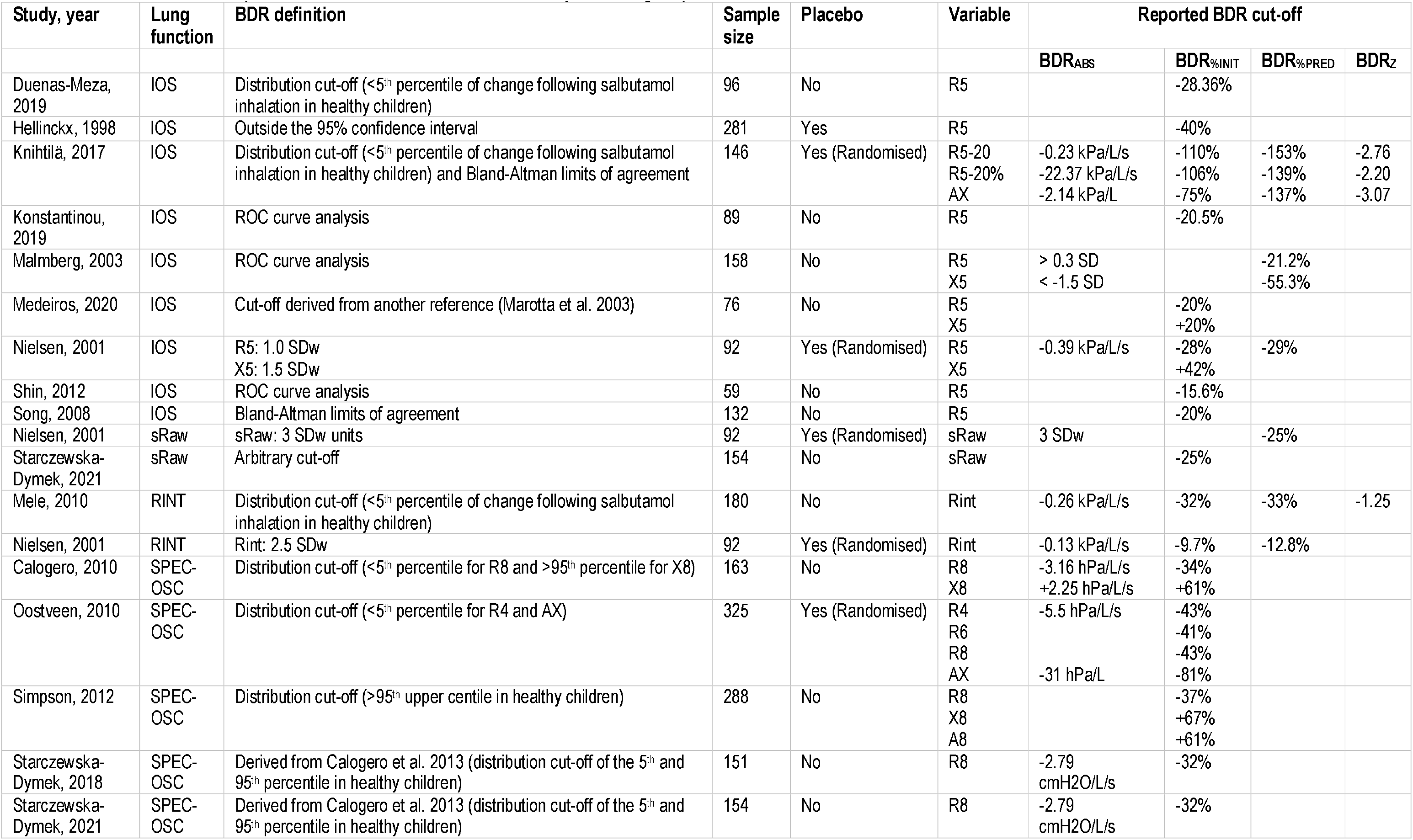
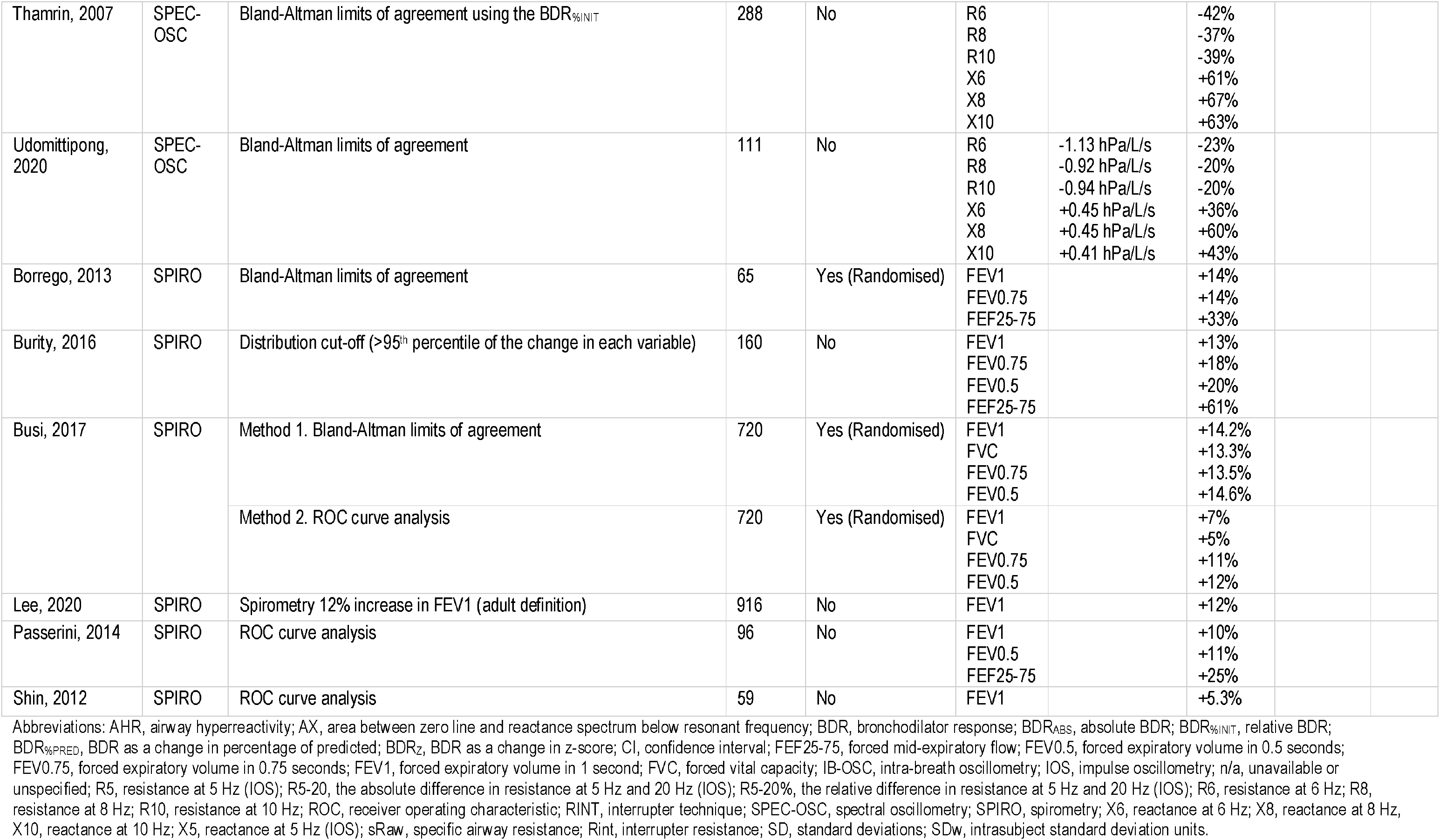
Bronchodilator response cut-offs in studies with a healthy control group.

**Figure 3.**
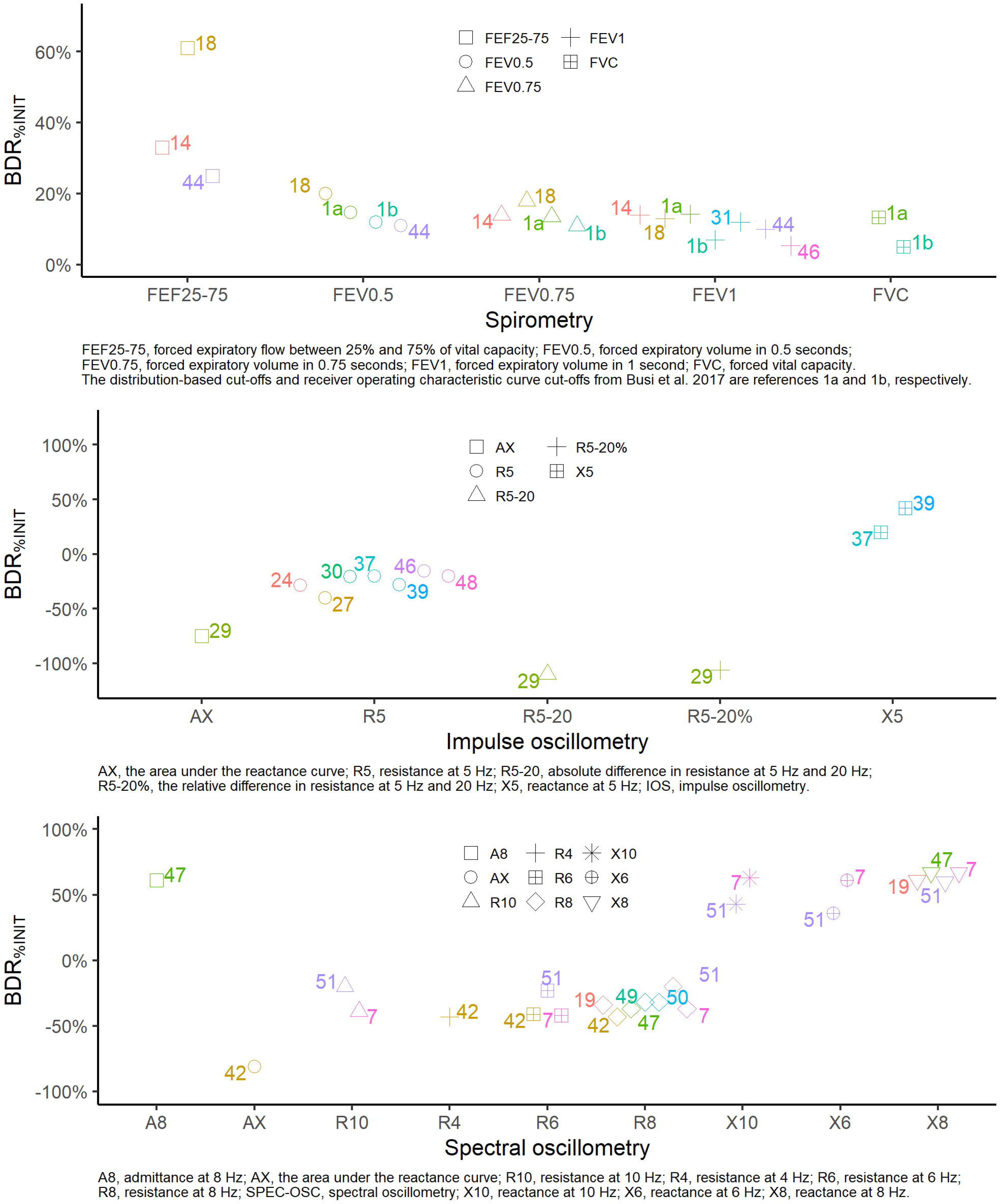
Studies reporting a bronchodilator response (BDR) as a percentage of the initial test (BDR_%INIT_) for spirometry (top), impulse oscillometry (middle), and spectral oscillometry (bottom) with shapes corresponding to lung function variables and colours corresponding to referenced studies.

### Spirometry

Of thirteen studies using spirometry, six (46.2%) incorporated a healthy control group and two (15.4%) included randomised placebo response testing (Table 2)^1,15,19,32,45,47^. Two studies used the within-subject between-test repeatability to define a BDR as the forced expiratory volume in one second (FEV_1_) +13.3-14% and forced expiratory volume in 0.75 seconds (FEV_0.75_) +13.5-14%. Three studies used receiver operating characteristic (ROC) curve analysis to define a BDR; one applied a distribution-based cut-off using >95^th^ percentile of the change in each variable in healthy children, and one study used an arbitrary cut-off of FEV_1_ +12%. All spirometry studies reported BDR_%INIT_, with none reporting BDR_ABS_ or BDR_%PRED_.

### Impulse oscillometry

Nine of fifteen (60%) studies using IOS included a healthy control group, with three incorporating placebo testing and all using the Jaeger MasterScreen IOS equipment (Table 2)^25,28,30,31,35,38,40,47,49^. The BDR_%INIT_ cut-off for R5 ranged from −15.6% to −40% and X5 from +20 to +42%. The only study in this review to report a BDR_Z_ was by Knihtila and colleagues, who reported cut-offs for R5-20 and AX^30^.

### Spectral oscillometry

Seven of eleven (63.6%) SPEC-OSC studies incorporated a healthy control group (Table 2)^8,20,43,48,50–52^. Of these studies, only Oostveen et al. 2010 included randomised, placebo response testing^43^. Four studies defined a BDR cut-off of >5^th^ percentile for resistance outcomes (R4, R6, R8, R10) and >95^th^ percentile in reactance outcomes (X6, X8, X10, AX). The BDR definitions used by Starczewska-Dymek and colleagues ^50,51^ were based on those developed by Calogero and colleagues^54^, who used a distribution-based cut-off of <5^th^ and >95^th^ percentile change following bronchodilator in healthy children. Udomittipong et al. defined a BDR using the within-subject between-test repeatability ^55^. All seven studies with healthy controls reported BDR_%INIT_, and five reported both BDR_ABS_ and BDR_%INIT_.^8,20,43,48,50–52^

### Interrupter technique

Two of the ten studies using the interrupter technique included a healthy control group (Table 2). Mele et al. defined a BDR cut-off as the <5^th^ percentile of change following salbutamol in 60 healthy children and reported BDR_ABS,_ BDR_%INIT,_ BDR_%PRED,_ and BDR_Z_^39^. Nielson et al. defined a BDR cut-off from 92 children as 2.5 intrasubject standard deviation units (SDw) and reported BDR_ABS,_ BDR_%INIT,_ and BDR_%PRED_^40^.

### sRaw

Two out of six sRaw studies included a healthy control group (Table 2)^40,51^. Only Nielson et al. 2001 derived a BDR_ABS_ cut-off from their study data for sRaw as a change of three SDw and BDR_%PRED_ as −25%^40^.

## DISCUSSION

The interest in and clinical utility of alternatives to spirometry for preschoolers is becoming more evident. Our review has revealed the need for greater standardisation of, and more consistency in, how a BDR assessment is conducted, defined, and reported across the literature. It is crucial to have a precise and feasible evaluation of lung function during preschool to enable early detection and intervention for airway disease, monitor longitudinal growth, and adjust for lung function evolution over this period^3^. Standardising BDR measurements across different techniques is a critical first step towards achieving this goal.

The highest feasibility was found with impulse and spectral oscillometry (Table 2). This is not surprising as oscillometry measures respiratory impedance during tidal breathing and requires no voluntary breathing manoeuvres. Spirometry had the lowest feasibility, reflecting the difficulty that preschoolers have generating sufficient force to achieve and maintain expiratory flow limitation to the point of residual volume. Only 41% of healthy children under four years old can produce an acceptable FEV_1_; most have a ratio of FEV_1_ to forced vital capacity (FVC) of greater than 90% due to the smaller lung volumes relative to airway calibre^56^. The use of preschool-relevant spirometry outcomes like the forced expiratory volume in 0.5 seconds (FEV_0.5_) and FEV_0.75_ would improve feasibility but have yet to be incorporated in the latest European Respiratory Society (ERS) and American Thoracic Society (ATS) technical standards due to a lack of systematic studies with these outcomes^6^.

Five studies did not specify whether a spacer delivered the bronchodilator. Although no significant differences were reported regarding spacer manufacturer, size, or use of facemasks versus mouthpieces for measuring a BDR using spirometry^57,58^, this has not been assessed for other lung function techniques in preschool-aged children. It is recommended that the spacer apparatus be specified for all studies.

Although the choice of bronchodilator and dosing depends on the clinical question to be answered, it is still an assumption that a clinically significant BDR is similar with SABA doses between 200-600 mcg^54,58^. The time between preBD and postBD testing ranged between 10-20 minutes, with seven studies having no mention in the methods of how long they waited between measurements. A minimum testing interval of 20 minutes is recommended for BDR testing in school-aged children using spirometry, but this has not been studied in preschoolers^58,59^. Our analysis highlights the need for further research comparing different bronchodilator protocols in identical subjects and has been echoed in a recent lung function standards document^6^.

It is also difficult to distinguish between biological or methodological effects on variability when dealing with small sample sizes, leading to sampling errors^60^. Quanjer et al. recommend sample sizes of more than 300 healthy subjects (150 females and 150 males) for generating spirometry reference values^60^. However, only half of the studies in this systematic review included a healthy control group, and only five accounted for the within-subject intrasession repeatability of the test using paired prePL and postPL testing. Of the studies reviewed, only two had a sample size greater than 300, included a healthy control group with randomisation for placebo response testing, and derived BDR cut-offs using within-session and between-session repeatability^1,43^. Data pooling for a more robust analysis is limited by the heterogeneity between studies in BDR methodology.

For preschool spirometry, Busi et al. and Borrego et al. accounted for the repeatability of testing in wheezy preschoolers and healthy controls; a preschool relative BDR of ≥12% for FEV_0.5_ and ≥11-14% for FEV_0.75_ may be appropriate^1,15^.

Despite all IOS studies utilising the same equipment (Jaeger MasterScreen IOS), a wide range of BDR_%INIT_ cut-offs for both R5 and X5 were reported, which may reflect heterogeneous methodologies to define BDR, population differences, poor reproducibility between laboratories, or all the above. The range of BDR_%INIT_ cut-offs for R8 and X8 was −20% to −43% and +60% to +67%, respectively (Table 2). Since a BDR_ABS_ will depend more on the preBD values, children with higher initial resistance (Rrs) or lower initial reactance (Xrs) will be more likely to reach a BDR_ABS_ cut-off and be diagnosed with reversible airway disease^54^.

Reporting BDR_%PRED_ or BDR_Z_ is ideal for all lung function modalities as it is population-specific and independent of subject age, height, and the magnitude of the preBD value^6^. Future studies involving preschool spirometry should incorporate BDR_%PRED_ or BDR_Z_ as there is spirometry reference data available for three years and older from the Global Lung Function Initiative (GLI). However, normative data are lacking for many techniques, especially for non-Caucasian children under six years old. Korea, Columbia, and Thailand were the only non-Caucasian predominant populations included in this review. There were no studies involving First Nations or African American children under six years old.

Our findings have significant implications for determining the clinical relevance of lung function changes following bronchodilator administration. To accurately assess the minimal clinically important difference (MCID) of BDR for diagnostic purposes, such as identifying asthma, and to evaluate the impact of interventions within clinical trials and individual young children, it is crucial to standardise preschool BDR methodology by establishing consensus definitions^61^.

For example, nearly half of all preschool-aged children worldwide experience asthma-like symptoms, but only 30% of those with recurrent wheezing will develop asthma beyond six years^62–64^. A distribution-based MCID, often defined as a change greater than the expected measurement error, can be calculated based on standardised response mean by taking the difference between preBD and postBD values and dividing by the standard deviation of the difference (SD_diff_­)^65–68^. This method relies heavily on sample sizes as SD_diff_ will be inversely proportional to the sample size. Early suggestions defining a distribution-based MCID as roughly equivalent to 0.5 SD_diff_ of the sample may only reflect the smallest detectable difference (SDD) for the test and may not be clinically perceivable by the clinician or patient^68^. Diagnosing asthma based on a positive BDR >0.5 SD in preschool wheezing may over-diagnose children with asthma and lead to unnecessary exposure to corticosteroids.

Some argue that the minimal detectable change (MDC) is a surrogate for an MCID, where the MDC based on the 95% confidence interval is 1.96 x √2 x SEM, where SEM is the standard error of the measurement and sample independent. Further studies are needed in children with disease (e.g., asthma) using an anchor-based MCID where the change in lung function is compared to the change in a clinical anchor (e.g., a patient’s reported symptoms).

This review included 43 studies and covered six established and emerging lung function testing modalities. To our knowledge, all studies followed recommended testing guidelines on equipment that met ERS/ATS requirements. Our quality assessment (Table S8) revealed significant heterogeneity among results from studies that used different lung function equipment, which could not be pooled for meta-analyses, even when controlling for methodology and outcome variables. Dandurand et al. demonstrated this comparability problem by showing significant differences in measurements of high-impedance test loads using IOS and SPEC-OSC devices^69^. Clinical and methodological diversity made pooling lung function data and meta-analyses impractical even for studies of the same lung function modality.

Several studies were excluded from this review as we could not separate preschool-aged data from larger cohort populations. Furthermore, we cannot comment on the potential learning effect sometimes experienced between preBD and postBD testing and acknowledge that this may create some bias in reported feasibility data. However, this is more likely to be experienced in techniques requiring specific manoeuvres (e.g., spirometry and sRaw), which scored lowest in overall feasibility. As previously mentioned, there was limited ethnic variation in our review, with only six studies performed in non-Caucasian predominant populations.

## CONCLUSION

Our review found significant inconsistencies in BDR assessment for preschool-aged children. This includes inconsistencies in testing methodology, cut-off determination, and BDR reporting. Defining a BDR in 2-6-year-old children is impossible based on available literature. Conventional techniques like spirometry and sRaw were less feasible for BDR testing in preschoolers. In contrast, tidal breathing-based techniques like oscillometry were easier to perform and can be incorporated more easily into clinical settings. Future research should aim to generate normative data, standardise BDR delivery, and consider the MCID in children with airway disease. The applicability of the Caucasian-derived estimate of BDR to diverse ethnic populations requires further clarification.

## Data Availability

The data are available from the corresponding author on reasonable request.

## REFERENCES

1. Busi LE, Restuccia S, Tourres R, Sly PD. Assessing bronchodilator response in preschool children using spirometry. Thorax 2017;72(4):367–372.

2. Pellegrino R, Antonelli A, Mondino M. Bronchodilator testing: an endless story. Eur Respir J 2010;35(5):952–4.

3. Beydon N, Davis SD, Lombardi E, Allen JL, Arets HG, Aurora P, Bisgaard H, Davis GM, Ducharme FM, Eigen H, et al. An official American Thoracic Society/European Respiratory Society statement: pulmonary function testing in preschool children. Am J Respir Crit Care Med 2007;175(12):1304–45.

4. Rosenfeld M, Allen J, Arets BH, Aurora P, Beydon N, Calogero C, Castile RG, Davis SD, Fuchs S, Gappa M, et al. An official American Thoracic Society workshop report: optimal lung function tests for monitoring cystic fibrosis, bronchopulmonary dysplasia, and recurrent wheezing in children less than 6 years of age. Ann Am Thorac Soc 2013;10(2):S1–S11.

5. Beydon N. Assessment of bronchial responsiveness in preschool children. Paediatric Respiratory Reviews 2006;7:S23–S25.

6. Stanojevic S, Kaminsky DA, Miller MR, Thompson B, Aliverti A, Barjaktarevic I, Cooper BG, Culver B, Derom E, Hall GL, et al. ERS/ATS technical standard on interpretive strategies for routine lung function tests. Eur Respir J 2022;60(1):2101499.

7. Thamrin C, Gangell CL, Kusel MMH, Schultz A, Hall GL, Stick SM, Sly PD. Expression of bronchodilator response using forced oscillation technique measurements: absolute versus relative. European Respiratory Journal 2010;36(1):212–212.

8. Thamrin C, Gangell CL, Udomittipong K, Kusel MM, Patterson H, Fukushima T, Schultz A, Hall GL, Stick SM, Sly PD. Assessment of bronchodilator responsiveness in preschool children using forced oscillations. Thorax 2007;62(9):814–9.

9. Bland JM, Altman DG. Statistical methods for assessing agreement between two methods of clinical measurement. Lancet 1986;1(8476):307–310.

10. Cotes JE. Absolute FEV1 values. Lancet 1981;2(8243):423.

11. Demedts M. Precise diagnosis of airflow obstruction - does it matter for treatment? The assessment of reversibility: what physiological tests? Eur Respir J 1990;3(9):1084–1087.

12. Munn Z, Moola S, Lisy K, Riitano D, Tufanaru C. Methodological guidance for systematic reviews of observational epidemiological studies reporting prevalence and cumulative incidence data. Int J Evid Based Healthc 2015;13(3):147–153.

13. Beydon N, M’Buila C, Peiffer C, Bernard A, Zaccaria I, Denjean A. Can bronchodilator response predict bronchial response to methacholine in preschool coughers? Pediatr Pulmonol 2008;43(8):815–21.

14. Bokov P, Jallouli-Masmoudi D, Amat F, Houdouin V, Delclaux C. Small airway dysfunction is an independent dimension of wheezing disease in preschool children. Pediatr Allergy Immunol 2021.

15. Borrego LM, Stocks J, Almeida I, Stanojevic S, Antunes J, Leiria-Pinto P, Rosado-Pinto JE, Hoo AF. Bronchodilator responsiveness using spirometry in healthy and asthmatic preschool children. Arch Dis Child 2013;98(2):112–7.

16. Bridge PD, Ranganathan S, McKenzie SA. Measurement of airway resistance using the interrupter technique in preschool children in the ambulatory setting. EUROPEAN RESPIRATORY JOURNAL 1999;13(4):792–796.

17. Bridge PD, McKenzie SA. Airway resistance measured by the interrupter technique: expiration or inspiration, mean or median? Eur Respir J 2001;17(3):495–8.

18. Bridge PD, Wertheim D, Jackson AC, McKenzie SA. Pressure oscillation amplitude after interruption of tidal breathing as an index of change in airway mechanics in preschool children. Pediatr Pulmonol 2005;40(5):420–5.

19. Burity EF, Pereira CA, Jones MH, Sayão LB, Andrade AD, Britto MC. Bronchodilator response cut-off points and FEV 0.75 reference values for spirometry in preschoolers. J Bras Pneumol 2016;42(5):326–332.

20. Calogero C, Parri N, Baccini A, Cuomo B, Palumbo M, Novembre E, Morello P, Azzari C, Martino M, Sly PD, et al. Respiratory impedance and bronchodilator response in healthy Italian preschool children. Pediatr Pulmonol 2010;45(11):1086–94.

21. Czovek D, Shackleton C, Hantos Z, Taylor K, Kumar A, Chacko A, Ware RS, Makan G, Radics B, Gingl Z, et al. Tidal changes in respiratory resistance are sensitive indicators of airway obstruction in children. THORAX 2016;71(10):907–915.

22. Da Silva Sena CR, Morten M, Meredith J, Kepreotes E, V EM, P GG, P DR, P DS, Whitehead B, Karmaus W, et al. Rhinovirus bronchiolitis, maternal asthma, and the development of asthma and lung function impairments. Pediatr Pulmonol 2021;56(2):362–370.

23. Devereux G, Turner SW, Craig LC, McNeill G, Martindale S, Harbour PJ, Helms PJ, Seaton A. Low maternal vitamin E intake during pregnancy is associated with asthma in 5-year-old children. Am J Respir Crit Care Med 2006;174(5):499–507.

24. Dom S, Weyler JJ, Droste JH, Hagendorens MM, Dierckx E, Bridts CH, De Backer W, Oostveen E. Determinants of baseline lung function and bronchodilator response in 4-year-old children. Eur Respir J 2014;44(2):371–81.

25. Duenas-Meza E, Correa E, López E, Morales JC, Aguirre-Franco CE, Morantes-Ariza CF, Granados CE, González-García M. Impulse oscillometry reference values and bronchodilator response in three- to five-year old children living at high altitude. J Asthma Allergy 2019;12:263–271.

26. Friedman NL, McDonough JM, Zhang X, Hysinger EB, Adams KM, Allen JL. Bronchodilator responsiveness assessed by forced oscillometry and multiple breath washout techniques in preschool children. Pediatr Investig 2018;2(2):83–89.

27. Harrison J, Gibson AM, Johnson K, Singh G, Skoric B, Ranganathan S. Lung function in preschool children with a history of wheezing measured by forced oscillation and plethysmographic specific airway resistance. Pediatr Pulmonol 2010;45(11):1049–56.

28. Hellinckx J, De Boeck K, Demedts M. No paradoxical bronchodilator response with forced oscillation technique in children with cystic fibrosis. Chest 1998;113(1):55–9.

29. Jerzyńska J, Janas A, Galica K, Stelmach W, Woicka-Kolejwa K, Stelmach I. Total specific airway resistance vs spirometry in asthma evaluation in children in a large real-life population. Ann Allergy Asthma Immunol 2015;115(4):272–6.

30. Knihtila H, Kotaniemi-Syrjanen A, Pelkonen AS, Kalliola S, Makela MJ, Malmberg LP. Small airway oscillometry indices: Repeatability and bronchodilator responsiveness in young children. Pediatr Pulmonol 2017;52(10):1260–1267.

31. Konstantinou GN, Papadopoulos NG, Manousakis E, Xepapadaki P. Virus-Induced Asthma/Wheeze in Preschool Children: Longitudinal Assessment of Airflow Limitation Using Impulse Oscillometry. J Clin Med 2019;8(9).

32. Lee DH, Kwon JW, Kim HY, Seo JH, Kim HB, Lee SY, Jang GC, Song DJ, Kim WK, Jung YH, et al. Asthma predictive index as a useful diagnostic tool in preschool children: a cross-sectional study in Korea. Clin Exp Pediatr 2020;63(3):104–109.

33. Leiria-Pinto P, Carreiro-Martins P, Peralta I, Marques J, Finelli E, Alves C, Belo J, Alves M, Papoila AL, Neuparth N. Factors associated with asthma control in 121 preschool children. J Investig Allergol Clin Immunol 2020:0.

34. Lezana V, Gajardo A, Bofill L, Gutierrez M, Mora S, Castro-Rodriguez JA. Airway tone dysfunction among pre-schoolers with positive asthma predictive index: A case-control study. Allergol Immunopathol (Madr) 2017;45(2):169–174.

35. Malmberg LP, Pelkonen AS, Haahtela T, Turpeinen M. Exhaled nitric oxide rather than lung function distinguishes preschool children with probable asthma. Thorax 2003;58(6):494–9.

36. Mauger-Hamel P, Du Boisbaudry C, Leon K, Alavi Z, Giroux-Metges MA. Relationship between baseline and post-bronchodilator interrupter resistance and specific airway resistance in preschool children. ANNALS OF ALLERGY ASTHMA & IMMUNOLOGY 2020;124(4):366–372.

37. McKenzie SA, Bridge PD, Healy MJ. Airway resistance and atopy in preschool children with wheeze and cough. Eur Respir J 2000;15(5):833–8.

38. Medeiros D, Castro P, Bianca ACD, Sarinho E, Araújo JF, Correia Junior M, Rizzo JA. Impulse oscillometry: pulmonary function assessment in preschool children. Expert Rev Respir Med 2020;14(12):1261–1266.

39. Mele L, Sly PD, Calogero C, Bernardini R, Novembre E, Azzari C, De Martino M, Lombardi E. Assessment and validation of bronchodilation using the interrupter technique in preschool children: Rint Bronchodilator Response in Preschool Children. Pediatr Pulmonol 2010;45(7):633–638.

40. Nielsen KG, Bisgaard H. Discriminative capacity of bronchodilator response measured with three different lung function techniques in asthmatic and healthy children aged 2 to 5 years. Am J Respir Crit Care Med 2001;164(4):554–9.

41. Oh MA, Shim JY, Jung YH, Seo JH, Young Kim H, Kwon JW, Kim BJ, Kim HB, Kim WK, Lee SY, et al. Fraction of exhaled nitric oxide and wheezing phenotypes in preschool children. Pediatr Pulmonol 2013;48(6):563–70.

42. Olaguíbel JM, Alvarez-Puebla MJ, Anda M, Gómez B, García BE, Tabar AI, Arroabarren E. Comparative analysis of the bronchodilator response measured by impulse oscillometry (IOS), spirometry and body plethysmography in asthmatic children. J Investig Allergol Clin Immunol 2005;15(2):102–6.

43. Oostveen E, Dom S, Desager K, Hagendorens M, De Backer W, Weyler J. Lung function and bronchodilator response in 4-year-old children with different wheezing phenotypes. Eur Respir J 2010;35(4):865–72.

44. Pao CS, Healy MJR, McKenzie SA. Airway resistance by the interrupter technique: Which algorithm for measuring pressure?? PEDIATRIC PULMONOLOGY 2004;37(1):31–36.

45. Passerini ML, Peirano RM, Estay IC, Becerra ID, Castro-Rodriguez JA. Utility of bronchodilator response for asthma diagnosis in Latino preschoolers. ALLERGOLOGIA ET IMMUNOPATHOLOGIA 2014;42(6):553–559.

46. Sarria EE, Mattiello R, Yao WG, Chakr V, Tiller CJ, Kisling J, Tabbey R, Yu ZS, Kaplan MH, Tepper RS. Atopy, Cytokine Production, and Airway Reactivity as Predictors of Pre-School Asthma and Airway Responsiveness. PEDIATRIC PULMONOLOGY 2014;49(2):132–139.

47. Shin YH, Jang SJ, Yoon JW, Jee HM, Choi SH, Yum HY, Han MY. Oscillometric and spirometric bronchodilator response in preschool children with and without asthma. Can Respir J 2012;19(4):273–7.

48. Simpson SJ, Straszek SP, Sly PD, Stick SM, Hall GL. Clinical investigation of respiratory system admittance in preschool children. Pediatr Pulmonol 2012;47(1):53–8.

49. Song TW, Kim KW, Kim ES, Park JW, Sohn MH, Kim KE. Utility of impulse oscillometry in young children with asthma. Pediatr Allergy Immunol 2008;19(8):763–8.

50. Starczewska-Dymeke L, Bozek A, Jakalski M. The Usefulness of the Forced Oscillation Technique in the Diagnosis of Bronchial Asthma in Children. CANADIAN RESPIRATORY JOURNAL 2018;2018.

51. Starczewska-Dymek L, Bozek A, Mielnik M. The sensitivity and specificity of the forced oscillation technique in the diagnosis of bronchoconstriction in children. JOURNAL OF ASTHMA 2021;58(3):334–339.

52. Udomittipong K, Triwatanawong J, Nimmannit A, Komoltri C. Cut-off values for positive bronchodilator response in healthy Thai preschool children using forced oscillation technique. Asian Pac J Allergy Immunol 2020;38(4):258–263.

53. Vilozni D, Barak A, Efrati O, Angarten A, Springer C, Yahav Y, Bentur L. The role of computer games in measuring spirometry in healthy and “asthmatic” preschool children. CHEST 2005;128(3):1146–1155.

54. Calogero C, Simpson SJ, Lombardi E, Parri N, Cuomo B, Palumbo M, de Martino M, Shackleton C, Verheggen M, Gavidia T, et al. Respiratory impedance and bronchodilator responsiveness in healthy children aged 2-13 years. Pediatr Pulmonol 2013;48(7):707–15.

55. Udomittipong K, Srisukhon W, Nimmannit A, Komoltri C. Respiratory Impedance Reference Values for Forced Oscillation Technique Predicted by Arm Span and Height in Thai Preschool Children. Pediatric allergy, immunology, and pulmonology 2017;30(2):97–102.

56. Aurora P, Stocks J, Oliver C, Saunders C, Castle R, Chaziparasidis G, Bush A. Quality Control for Spirometry in Preschool Children with and without Lung Disease. Am J Respir Crit Care Med 2004;169(10):1152–1159.

57. Zar HJ. Lung deposition of aerosol---a comparison of different spacers. Archives of Disease in Childhood 2000;82(6):495–498.

58. D’Vaz N, Okitika TA, Shackleton C, Devadason SG, Hall GL. Bronchodilator responsiveness in children with asthma is not influenced by spacer device selection. Pediatr Pulmonol 2019;54(5):531–536.

59. Stavreska V, Verheggen M, Oostryck J, Stick SM, Hall GL. Determining the time to maximal bronchodilator response in asthmatic children. J Asthma 2009;46(1):25–29.

60. Quanjer PH, Stocks J, Cole TJ, Hall GL, Stanojevic S, on behalf of the Global Lungs Initiative. Influence of secular trends and sample size on reference equations for lung function tests. European Respiratory Journal 2011;37(3):658–664.

61. Waalkens HJ, Merkus PJ, van Essen-Zandvliet EE, Brand PL, Gerritsen J, Duiverman EJ, Kerrebijn KF, Knol KK, Quanjer PH. Assessment of bronchodilator response in children with asthma. Dutch CNSLD Study Group. Eur Respir J 1993;6(5):645–651.

62. Martinez FD, Wright AL, Taussig LM, Holberg CJ, Halonen M, Morgan WJ. Asthma and wheezing in the first six years of life. The Group Health Medical Associates. N Engl J Med 1995;332(3):133–8.

63. Bisgaard H. Use of inhaled corticosteroids in pediatric asthma. Pediatr Pulmonol Suppl 1997;15:27–33.

64. Castro-Rodriguez JA. The necessity of having asthma predictive scores in children. J Allergy Clin Immunol 2013;132(6):1311–3.

65. Draak THP, de Greef BTA, Faber CG, Merkies ISJ, PeriNomS study group. The minimum clinically important difference: which direction to take. Eur J Neurol 2019;26(6):850–855.

66. Norman GR, Sloan JA, Wyrwich KW. Interpretation of changes in health-related quality of life: the remarkable universality of half a standard deviation. Med Care 2003;41(5):582–592.

67. Jones PW, Beeh KM, Chapman KR, Decramer M, Mahler DA, Wedzicha JA. Minimal clinically important differences in pharmacological trials. Am J Respir Crit Care Med 2014;189(3):250–5.

68. Woaye-Hune P, Hardouin J-B, Lehur P-A, Meurette G, Vanier A. Practical issues encountered while determining Minimal Clinically Important Difference in Patient-Reported Outcomes. Health Qual Life Outcomes 2020;18(1):156.

69. Dandurand RJ, Lavoie JP, Lands LC, Hantos Z, Oscillometry Harmonisation Study G. Comparison of oscillometry devices using active mechanical test loads. ERJ Open Res 2019;5(4).

