## Supplementary material for "Assessment of a bronchodilator response in preschoolers: a systematic review": Table S1

Table S1. Database search strategies

PUBMED = 367

((oscillometry[tiab] OR "forced oscillation technique"[tiab] OR "oscillation"[tiab] OR "FOT"[tiab] OR "oscillations"[tiab] OR “Oscillometry”[Mesh] OR "interrupter technique"[tiab] OR "plethysmography"[tiab] OR “spirometry”[tiab]) AND (child*[tiab] OR toddler*[tiab] OR infant*[tiab] OR baby[tiab] OR babies[tiab] OR pediatric*[tiab] OR paediatric*[tiab] OR "Child"[Mesh] OR "Child, Preschool"[Mesh] OR "Infant"[Mesh]) AND (bronchodilator[tiab]) AND (lung*[tiab] OR respiratory[tiab] OR pulmonary[tiab] OR "Lung"[Mesh] OR "Respiration"[Mesh]))

EMBASE = 670

((oscillometry:ti,ab OR "forced oscillation technique":ti,ab OR "oscillation":ti,ab OR "FOT":ti,ab OR "oscillations":ti,ab OR “Oscillometry”/de OR "interrupter technique":ti,ab OR "plethysmography":ti,ab OR “spirometry”:ti,ab) AND (child*:ti,ab OR toddler*:ti,ab OR infant*:ti,ab OR baby:ti,ab OR babies:ti,ab OR pediatric*:ti,ab OR paediatric*:ti,ab OR "Child"/de OR "Child, Preschool"/de OR "Infant"/de) AND (bronchodilator:ti,ab) AND (lung*:ti,ab OR respiratory:ti,ab OR pulmonary:ti,ab OR "Lung"/de OR "Respiration"/de) AND [embase]/lim)

CINAHL = 86

((TI oscillometry OR AB oscillometry OR TI "forced oscillation technique" or AB “forced oscillation technique” OR TI "oscillation" OR AB “oscillation” OR TI "FOT" or AB “FOT” OR TI "oscillations" or AB “oscillations” OR MH “Oscillometry” OR TI "interrupter technique" OR AB “interrupter technique” OR TI "plethysmography" OR AB “plethysmography” OR TI “spirometry” OR AB “spirometry”) AND (TI child* OR AB child* OR TI toddler* OR AB toddler* OR TI infant* OR AB infant* OR TI baby OR AB baby OR TI babies OR AB babies OR TI pediatric* OR AB pediatric* OR TI paediatric* OR AB paediatric* OR MH "Child" OR MH "Preschool" OR MH "Infant") AND (TI bronchodilator OR AB bronchodilator) AND (TI lung* OR AB lung OR TI respiratory OR AB respiratory OR TI pulmonary OR AB pulmonary OR MH "Lung" OR MH "Respiration"))

Web of Science = 649

((oscillometry) OR ("forced oscillation technique") OR ("oscillation") OR ("FOT") OR ("oscillations") OR (“Oscillometry”) OR ("interrupter technique") OR ("plethysmography") OR (“spirometry”)) AND (child* OR toddler* OR infant* OR baby OR babies OR pediatric* OR paediatric* OR "Child" OR "Child, Preschool" OR "Infant") AND bronchodilator AND (lung* OR respiratory OR pulmonary OR "Lung" OR "Respiration")
