## Supplementary material for "Assessment of a bronchodilator response in preschoolers: a systematic review": Table S2

Table S2. Data extraction variables from each lung function technique

| Lung Function Technique | Lung Function Variables Extracted* |
| --- | --- |
| Spirometry | Forced expiratory volume in one second (FEV_1_)  Forced expiratory volume in 0.5 seconds (FEV_0.5_)  Forced expiratory volume in 0.75 seconds (FEV_0.75_)  Forced vital capacity (FVC)  Forced expiratory flow between 25% and 75% of vital capacity (FEF_25-75_) |
| Impulse oscillometry | Resistance at 5 Hz (R5)  Resistance at 10 Hz (R10)  Resistance at 15 Hz (R15)  Resistance at 20 Hz (R20)  Resistance at 25 Hz (R25)  Resistance at 35 Hz (R35)  Resistance between 5-20 Hz (R5-20)  Reactance at 5 Hz (X5)  Reactance at 10 Hz (X10)  Reactance at 15 Hz (X15)  Reactance at 20 Hz (X20)  Reactance at 25 Hz (X25)  Reactance at 35 Hz (X35)  Resonant frequency (Fres)  Area under the reactance curve to resonant frequency (AX) |
| Spectral oscillometry | Resistance at 4 Hz (R4)  Resistance at 6 Hz (R6)  Resistance at 8 Hz (R8)  Resistance at 10 Hz (R10)  Resistance at 12 Hz (R12)  Reactance at 4 Hz (X4)  Reactance at 6 Hz (X6)  Reactance at 8 Hz (X8)  Reactance at 10 Hz (X10)  Reactance at 12 Hz (X12) |
| Intra-breath oscillometry | Resistance at end-expiration at 10 Hz (ReE)  Resistance at end-inspiration at 10 Hz (ReI)  Tidal change in resistance at 10 Hz (ΔR = ReE – ReI)  Reactance at end-expiration at 10 Hz (XeE)  Reactance at end-inspiration at 10 Hz (ReI)  Tidal change in reactance at 10 Hz (ΔX = XeE – XeI) |
| Interrupter technique | Interrupter resistance (Rint) |
| Specific airway resistance | Specific airway resistance (sRaw) |

* Includes pre-/post-bronchodilator and pre-/post-placebo measurements
