## Supplementary material for "Assessment of a bronchodilator response in preschoolers: a systematic review": Table S3

Table S3. Spirometry bronchodilator response in preschool-aged children

| **Study, year** | **Lung function equipment** | **Sample size  n (F:M)** | **Acceptable BDR (feasibility %)** | **Bronchodilator dose** | **Spacer** | **Time (min)** | **Variables with derived BDR**  **cut-off** |
| --- | --- | --- | --- | --- | --- | --- | --- |
| Borrego, 2013 | MasterScreen Body (V.4.6.5, Jaeger, San Diego, USA) | HL 22 (12:10) WZ 43 (15:28) | HL 22 of 25 (88%) WZ 43 of 43 (100%) | Salbutamol 400 mcg | Yes | n/a | FEV1, FEV0.75, FEV25-75 |
| Burity, 2016 | Portal spirometer validated by the ATS (Koko; Ferraris Respiratory, Louisville, CO, USA) | HL 160 (76:84) | 160 of 216 (74.1%) | Albuterol 400 mcg | Yes | 15 | FEV1, FEV0.5, FEV0.75, FEV25-75 |
| Busi, 2016 | n/a | HL 431 (211:220) WZ 289 (118:171) | HL 320 of 364 (87.9%)* WZ 233 of 244 (95.5%)* | Salbutamol 400 mcg | Yes | 15 | FVC, FEV1, FEV0.5, FEV0.75 |
| Devereux, 2006 | Spirotrac IV version 4.22; Vitalograph, Maids Moreton, UK) with onscreen incentive software. | Birth cohort 502 (n/a) | 269 of 502 (53.6%)** | Albuterol 400 mcg | Yes | 15 | None |
| Jerzyńska, 2015 | MasterScreen (Erich Jaeger GmbH, Hochberg, Germany) | 142 (n/a) | 788 of 885 (89%)*** | Salbutamol 200 mcg | Yes | 15 | None |
| Lee, 2020 | VMAX 22 (Sensormedics, Anaheim, CA, USA) | HL 880 (431:449) WZ 36 (15:21) | HL 466 of 880 (53%) WZ 16 of 36 (44.4%) | Salbutamol 200 mcg | n/a | 15 | None |
| Leiria-Pinto, 2020 | MasterScreen (v4.6., Jaeger Co) using ATS guidelines | HL 15 (7:7) WZ 107 (46:61) | HL 15 of 17 (88.2%) WZ 114 of 139 (82%) | Salbutamol 400 mcg | Yes | 15 | None |
| Olaguíbel, 2005 | MasterScreen (Jaeger, Med Point Technologies, Inc., Milbury, OH) | WZ 36 (11:25) | 28 of 33 (84.8%) | Salbutamol 400 mcg | Yes | 15 | None |
| Passerini, 2014 | MasterScreen (Jaeger, Wurzburg, Germany | HL 32 (19:13) WZ 64 (35:29) | HL 30 of 32 (93.8%) WZ 61 of 64 (95.3%) | Salbutamol 200 mcg | Yes | n/a | FEV1, FEV0.5, FEF25-75 |
| Sarria, 2014 | n/a | 74 (n/a) | 54 of 74 (73%) | Albuterol 500 mcg | n/a | n/a | None |
| Shin, 2012 | MasterScreen (Jaeger, Wurzburg, Germany) | HL 29 (15:14) WZ 30 (16:14) | HL 29 of 29 (100%) WZ 30 of 30 (100%) | Salbutamol 400 mcg | Yes | 15 | FEV1 |
| Song, 2008 | MasterScreen (Jaeger, Wurzburg, Germany) | HL 55 (20:35) WZ 77 (29:48) | HL 55 of 55 (100%) WZ 77 of 77 (100%) | Salbutamol 200 mcg | Yes | 15 | None |
| Vilozni, 2005 | ZAN100; ZAN Messgeraete GmbH; Oberthulba, Germany | HL 109 (59:50) WZ 156 (62:94) | HL no BDR testing WZ 98 of 98 (100%) | Salbutamol 200 mcg | Yes | n/a | None |

Abbreviations:

* Feasibility calculated based on those children able to perform acceptable baseline spirometry.

** Feasibility calculated based on acceptable paired pre-/post-bronchodilator measurements out of total sample size.

*** Feasibility for ages 4-18 (no sub-analysis for preschool ages).

† BDR cut-off from Miller et al. Eur Respir J 2005.

FEF25-75, forced mid-expiratory flow; FEV0.5, forced expiratory volume in 0.5 seconds; FEV0.75, forced expiratory volume in 0.75 seconds; FEV1, forced expiratory volume in 1 second; FVC, forced vital capacity; HL, healthy; n/a, unavailable or unspecified; WZ, wheezy.
