## Supplementary material for "Assessment of a bronchodilator response in preschoolers: a systematic review": Table S4

Table S4. Impulse oscillometry bronchodilator response in preschool-aged children

| **Study, year** | **Lung function equipment** | **Sample size**  **n (F:M)** | **Acceptable BDR**  **(feasibility %)** | **Bronchodilator dose** | **Spacer** | **Time** | **BDR cut-off** | **Variables with derived BDR**  **cut-off** |
| --- | --- | --- | --- | --- | --- | --- | --- | --- |
| Bokov, 2021 | MasterScope Body  (CareFusion Technologies, USA) | WZ: 139 (45:94) | WZ: 139 of 160 (86.9%) | Salbutamol 400 mcg | n/a | n/a | No |  |
| Da Silva Sena, 2021 | MasterScreen IOS  (Jaeger, Wurzburg, Germany) | Past bronchiolitis:  139 (49:90) | 104 of 107 (97.2%) | Salbutamol 400 mcg | Yes | 15 min | Yes | R5 |
| Duenas-Meza, 2019 | MasterScreen IOS | HL: 96 (58:38) | 96 of 96 (100%) | Salbutamol 400 mcg | Yes | 15 min | Yes | R5 |
| Hellinckx, 1998 | MasterScreen IOS | HL and WZ: 281 (152:129) | 281 of 337 (83.4%) | Salbutamol 200 mcg | Yes | 20 min | Yes | R5 |
| Knihtilä, 2017 | MasterScreen IOS | HL: 103 (50:53) WZ: 43 (12:31) | HL: 103 of 103 (100%) WZ: 43 of 43 (100%) | Salbutamol 300 mcg | Yes | 15 min | Yes | R5-20  AX |
| Konstantinou, 2019 | MasterScreen Spirometry-IOS (Jaeger) | HL: 46 (26:20) WZ: 43 (20:23) | 93 of 93 (100%) | Albuterol 400 mcg | Yes | 15 min | Yes | R5 |
| Leiria-Pinto, 2020 | MasterScreen Spirometry-IOS | HL: 14 (7:7) WZ: 107 (46:61) | 121 of 129 (93.8%) | Salbutamol 400 mcg | Yes | 15 min | No |  |
| Lezana, 2017 | MasterScreen Spirometry-IOS | WZ: 108 (52:56) | 108 of 109 (99.1%) | Salbutamol 200 mcg | Yes | 15 min | No |  |
| Malmberg, 2003 | MasterScreen IOS | HL: 62 (32:30) ASTH and cough: 96 (40:56) | 137 of 143 (95.8%) | Salbutamol 300 mcg | Yes | 15 min | Yes | R5  X5 |
| Medeiros, 2020 | MasterScreen IOS  (VIASYS Healthcare GmbH, Germany) | HL: 21 (10:11) WZ: 55 (35:20) | HL: 21 of 21 (100%) WZ: 55 of 55 (100%) | Salbutamol 200 mcg | Yes | 15 min | Yes | R5  X5 |
| Nielsen, 2001 | MasterScreen IOS and body (Jaeger) | HL: 37 (19:18)  WZ: 55 (25:30) | HL: 37 of 41 (90.2%) WZ: 55 of 55 (100%) | Terbutaline 500 mcg | Yes | 20 min | Yes | R5  X5 |
| Oh, 2013 | MasterScreen Spirometry-IOS | HL: 282 (144:138)  Early-onset WZ: 23 (10:13)  Late-onset WZ: 67 (34:33) | 273 of 372 (73.4%) | Salbutamol 200 mcg | n/a | 15 min | No |  |
| Olaguíbel, 2005 | MasterScreen System  (Jaeger, Milbury, OH) | WZ: 36 (11:25) | 33 of 36 (91.7%) | Salbutamol 400 mcg | Yes | 15 min | No |  |
| Shin, 2012 | MasterScreen Spirometry-IOS | HL: 29 (15:14) WZ: 30 (16:14) | HL and WZ: 59 of 59 (100%) | Salbutamol 400 mcg | Yes | 15 min | Yes | R5 |
| Song, 2008 | MasterScreen Spirometry-IOS | HL: 55 (20:35) WZ: 77 (29:48) | HL: 55 of 55 (100%) WZ: 77 of 77 (100%) | Salbutamol 200 mcg | Yes | 15 min | Yes | R5 |

Abbreviations:

ASTH, asthma; AX, the area between zero line and reactance spectrum below resonant frequency; BDR, bronchodilator response; HL, healthy; n/a, unavailable or unspecified; R5, resistance at 5 Hz; R5-20, the difference in resistance at 5 Hz and 20 Hz; SD, standard deviations; WZ, wheeze; X5, reactance at 5 Hz; Zrs, respiratory impedance.
