## Supplementary material for "Assessment of a bronchodilator response in preschoolers: a systematic review": Table S5

Table S5. Spectral and intra-breath oscillometry bronchodilator response in preschool-aged children.

| **Study, year** | **Lung function equipment** | **Lung function** | **Sample size**  **n (F:M)** | **Acceptable BDR (feasibility %)** | **Bronchodilator dose** | **Spacer** | **Time (min)** | **BDR cut-off** | **Variables with derived BDR**  **cut-off** |
| --- | --- | --- | --- | --- | --- | --- | --- | --- | --- |
| Calogero, 2010 | Quark i2m (COSMED, Rome, Italy) | SPEC-OSC | HL 163 (82:81) | 154 of 163 (94.5%) | Salbutamol 200 mcg | Yes | 15 | Yes | R8, X8 |
| Czovek, 2016* | Custom-made device | IB-OSC and SPEC-OSC | HL 23 WZ 17 | HL 23 of 23 (100%) WZ 17 of 20 (85%) | Salbutamol 400 mcg | Yes | n/a | No |  |
| Dom, 2014* | Custom-made device | SPEC-OSC | HL 535 (265:270) | HL 498 of 535 (93.1%) | Salbutamol 200 mcg | Yes | 15 | No |  |
| Friedman, 2018* | Quark i2m (COSMED, Rome, Italy) | SPEC-OSC | HL 28 (19:9) WZ 23 (12:11) | HL 27 of 28 (96.4%) WZ 22 of 23 (95.7%) | Albuterol 200 mcg | Yes | 10 | No |  |
| Harrison, 2010* | Quark i2m (COSMED, Rome, Italy) | SPEC-OSC | HL 24 (14:10) WZ 59 (24:35) | HL 23 of 24 (95.8%) WZ 54 of 59 (91.5%) | Salbutamol 400 mcg | Yes | 15 | No |  |
| Oostveen, 2010 | Custom-made device | SPEC-OSC | HL 144 (73:71) WZ 181 (79:102) | Total 313 of 325 (96.3%) | Salbutamol 200 mcg | Yes | 15 | Yes | R4, R6, R8, AX |
| Simpson, 2012^‡^ | i2m, Chess Medical, Belgium | SPEC-OSC | HL 78 (42:36) WZ 66 (25:41) ASTH 56 (21:35) CF 39 (24:15)  CNLD 49 (26:23) | Total 288 of 288 (100%) | Salbutamol 600 mcg | Yes | 15 | Yes | R8, X8, A8 |
| Starczewska-Dymek, 2021^†^ | Resmon Pro (Restech SRL, Italy) | SPEC-OSC | HL 52 (27:25)  WZ 102 (49:53) | Total 154 of 154 (100%) | Salbutamol 200 mcg | Yes | 15 | Yes | R8 |
| Starczewska-Dymek, 2018^†^ | Resmon Pro (Restech SRL, Italy) | SPEC-OSC | HL 45 (25:20)  WZ 53 (28:25)  ASTH, CF, CNLD 53 (26:27) | Total 151 of 151 (100%) | Salbutamol 200 mcg | Yes | 15 | Yes | R8 |
| Thamrin, 2007^‡^ | i2m, Chess Medical, Belgium | SPEC-OSC | HL 78 (42:36) WZ 66 (25:41) ASTH 56 (21:35) CF 39 (24:15) CNLD 49 (26:23) | Total 288 of 288 (100%) | Salbutamol 600 mcg | Yes | 15 | Yes | R6, R8, R10, X6, X8, X10 |
| Udomittipong, 2020 | Quark i2m (COSMED, Rome, Italy) | SPEC-OSC | HL 111 (60:51) | HL 111 of 150 (74%) | Salbutamol 400 mcg | Yes | 15-20 | Yes | R6, R8, R10, X6, X8, X10 |

Abbreviations: * No BDR cut-off data; † BDR cut-off data derived from Hellinckx et al. 1998 and not study data; ‡ same BDR data reported.

A8, admittance at 8 Hz; ASTH, asthma; AX, the area between zero line and reactance spectrum below resonant frequency; BDR, bronchodilator response; CF, cystic fibrosis; CNLD, chronic neonatal lung disease; HL, healthy; IB-OSC, intra-breath oscillometry; n/a, unavailable or unspecified; R6, resistance at 6 Hz; R8, resistance at 8 Hz; R10, resistance at 10 Hz; SD, standard deviations; SPEC-OSC, spectral oscillometry; WZ, wheeze; X6, reactance at 6 Hz; X8, reactance at 8 Hz, X10, reactance at 10 Hz; Zrs, respiratory impedance.
