## Supplementary material for "Assessment of a bronchodilator response in preschoolers: a systematic review": Table S6

Table S6. Interrupter technique bronchodilator response in preschool-aged children.

| **Study, year** | **Lung function equipment** | **Sample size**  **n (F:M)** | **Acceptable BDR**  **(feasibility %)** | **Bronchodilator dose** | **Spacer** | **Time (min)** | **BDR cut-off** |
| --- | --- | --- | --- | --- | --- | --- | --- |
| Beydon, 2008 | Spiroteq apparatus (Dyn'R Ltd, Toulouse, France) | Chronic cough but no WZ or ASTH 38 (23:15) | 38 of 38 (100%) | Salbutamol 400 mcg | Yes | 15 | Yes |
| Bokov, 2021 | SpiroDyn'R apparatus (Dyn'R Ltd, Toulouse, France) | WZ 139 (45:94) | 139 of 160 (86.9%) | Salbutamol 400 mcg | n/a | n/a | No |
| Bridge, 1999 | Microlab 4000 (Micromedical Ltd, Gillingham, UK) | Past WZ 32 (n/a) Active WZ 16 (n/a) | 2-3 years old 42 of 79 (53.2%) 3-4 years old 74 of 104 (71.2%) 4-5 years old 80 of 88 (90.9%) | Salbutamol 400 mcg | Yes | 15 | Yes |
| Bridge, 2001 | Microlab 4000 (Micromedical Ltd, Gillingham, UK) | Asymptomatic children with a history of respiratory symptoms 40 (n/a) | 40 of 40 (100%) | Salbutamol 400 mcg | Yes | 15 | No |
| Bridge, 2005 | MicroRint (Micromedical, Rochester, UK) | WZ 25 (15:10) | 25 of 25 (100%) | Salbutamol 400 mcg | Yes | 15 | No |
| Mauger-Hamel, 2020 | SpiroDyn’R apparatus (3.2.0.5 version, Ltd, Toulouse, France) | WZ 130 (55:75) | 83 of 101 (82.2%) | Salbutamol 400 mcg | Yes | 15 | Yes |
| McKenzie, 2000 | Microlab 4000 (Micromedical Ltd, Gillingham, UK | HL 63 (33:30) WZ 82 (31:51)  Recurrent cough 58 (29:29) | HL 48 of 48 (100%)* WZ 82 of 82 (100%) Recurrent cough 58 of 58 (100%) | Salbutamol 400 mcg | Yes | n/a | No |
| Mele, 2010 | MicroRint (Micromedical, Rochester, UK) | HL 60 (23:37)  WZ asymptomatic 60 (24:36)  WZ symptomatic 60 (29:31) | 180 of 180 (100%) | Salbutamol 200 mcg | Yes | 15 | Yes |
| Nielsen, 2001 | MasterScreen (Erich Jaeger GmbH, Wurzburg, Germany) | HL 37 (19:18)  WZ 55 (25:30) | HL 37 of 41 (90.2%) WZ 55 of 55 (100%) | Terbutaline 500 mcg | Yes | 20 | Yes |
| Pao, 2004 | MicroRint (Micromedical) | WZ 39 (18:21) | 39 of 39 (100%) | Salbutamol 400 mcg | n/a | 20 | No |

Abbreviations:

* BDR is defined as the logarithm of the ratio of the pre-bronchodilator and post-bronchodilator tests.

ASTH, asthma; BDR, bronchodilator response; HL, healthy; n/a, unavailable or unspecified; Rint, interrupter resistance; WZ, wheeze.
