## Supplementary material for "Assessment of a bronchodilator response in preschoolers: a systematic review": Table S7

Table S7. Specific airway resistance bronchodilator response in preschool-aged children

| **Study, year** | **Lung function equipment** | **Sample size**  **n (F:M)** | **Acceptable BDR (feasibility %)** | **Bronchodilator dose** | **Spacer** | **Time (min)** | **BDR cut-off** |
| --- | --- | --- | --- | --- | --- | --- | --- |
| Harrison, 2010 | MasterScreen (Jaeger) | HL 24 (14:10) WZ 59 (24:35) | HL 20 of 24 (83.3%) WZ 36 of 59 (61%) | Salbutamol 400 mcg | Yes | 15 | No |
| Jerzyńska, 2015 | MasterScreen (Jaeger) | Asthma-like symptoms 142 (n/a) | 788 of 885 (89%) | Salbutamol 200 mcg | Yes | 15 | Yes |
| Mauger-Hamel, 2020 | MasterScreen (Jaeger) | WZ 130 (55:75) | 83 of 101 (82.2%) | Salbutamol 400 mcg | Yes | 15 | Yes |
| Nielsen, 2001 | MasterScreen (Jaeger) | HL 37 (19:18)  WZ 55 (25:30) | HL 37 of 41 (90.2%) WZ 55 of 55 (100%) | Terbutaline 500 mcg | Yes | 20 | Yes |
| Olaguíbel, 2005* | MasterScreen (Jaeger) | WZ 36 (11:25) | 36 of 36 (100%) | Salbutamol 400 mcg | Yes | 15 | Yes |
| Starczewska-Dymek, 2021 | n/a | HL 52 (27:25)  WZ 102 (49:53) | Total 154 of 154 (100%) | Salbutamol 200 mcg | Yes | 15 | Yes |

Abbreviations:

* The suggested BDR cut-off was derived from Ortiz et al. 2022 and not from study data.

BDR, bronchodilator response; CoV, coefficient of variation; HL, healthy; n/a, unavailable or unspecified; SD, standard deviations; sRaw, specific airway resistance; WZ, wheeze.
