## Supplementary material for "Assessment of a bronchodilator response in preschoolers: a systematic review": Table S8

Table S8. Joanna Briggs Institute (JBI) critical appraisal for included studies.

|  | Appropriate sample frame (healthy cohort)? | Study participants randomly sampled? | Sample size adequate (≥300 healthy children)? | Study subjects and setting described in detail? | Data analysis with sufficient coverage of the identified sample? | Did they account for the short-term repeatability of the test? | BDR measured in a standard, reliable way for all participants? | Appropriate statistical analysis? | Adequate response rate and management of low responses? |
| --- | --- | --- | --- | --- | --- | --- | --- | --- | --- |
| Beydon, 2008 | **N** | **N** | **N** | **Y** | **Y** | **N** | **Y** | **Y** | **Y** |
| Bokov, 2021 | **N** | **Y** | **N** | **Y** | **N** | **N** | **N** | **N** | **Y** |
| Borrego, 2013 | **Y** | **Y** | **N** | **Y** | **Y** | **Y** | **N** | **Y** | **Y** |
| Bridge, 1999 | **N** | **Y** | **N** | **N** | **N** | **Y** | **Y** | **Y** | **Y** |
| Bridge, 2001 | **N** | **N** | **N** | **N** | **N** | **N** | **N** | **N** | **N** |
| Bridge, 2005 | **N** | **N** | **N** | **N** | **Y** | **N** | **N** | **N** | **N** |
| Burity, 2016 | **Y** | **Y** | **N** | **Y** | **Y** | **N** | **Y** | **Y** | **Y** |
| Busi, 2017 | **Y** | **Y** | **Y** | **Y** | **Y** | **Y** | **Y** | **Y** | **Y** |
| Calogero, 2010 | **Y** | **Y** | **N** | **Y** | **Y** | **Y** | **Y** | **Y** | **Y** |
| Czӧvek, 2016 | **Y** | **Y** | **N** | **Y** | **N** | **N** | **N** | **N** | **Y** |
| Da Silva Sena, 2021 | **N** | **N** | **N** | **Y** | **N** | **N** | **Y** | **Y** | **Y** |
| Devereux, 2006 | **N** | **Y** | **Y** | **N** | **N** | **N** | **N** | **N** | **Y** |
| Dom, 2014 | **Y** | **Y** | **Y** | **Y** | **N** | **N** | **N** | **N** | **Y** |
| Duenas-Meza, 2019 | **Y** | **Y** | **N** | **Y** | **Y** | **N** | **Y** | **Y** | **Y** |
| Friedman, 2018 | **Y** | **Y** | **N** | **Y** | **Y** | **N** | **N** | **N** | **Y** |
| Harrison, 2010 | **Y** | **Y** | **N** | **Y** | **Y** | **N** | **N** | **N** | **Y** |
| Hellinckx, 1998 | **Y** | **Y** | **N** | **Y** | **?** | **Y** | **Y** | **Y** | **Y** |
| Jerzyńska, 2015 | **N** | **Y** | **N** | **N** | **N** | **N** | **Y** | **Y** | **Y** |
| Knihtilä, 2017 | **Y** | **Y** | **N** | **Y** | **Y** | **Y** | **Y** | **Y** | **N** |
| Konstantinou, 2019 | **Y** | **Y** | **N** | **Y** | **Y** | **N** | **Y** | **Y** | **Y** |
| Lee, 2020 | **Y** | **Y** | **Y** | **Y** | **Y** | **N** | **N** | **N** | **N** |
| Leiria-Pinto, 2020 | **Y** | **Y** | **N** | **Y** | **Y** | **N** | **N** | **N** | **Y** |
| Lezana, 2017 | **N** | **Y** | **N** | **Y** | **Y** | **N** | **N** | **N** | **Y** |
| Malmberg, 2003 | **Y** | **Y** | **N** | **Y** | **Y** | **N** | **Y** | **Y** | **Y** |
| Mauger-Hamel, 2020 | **N** | **Y** | **N** | **Y** | **Y** | **N** | **Y** | **Y** | **Y** |
| McKenzie, 2000 | **Y** | **Y** | **N** | **Y** | **Y** | **N** | **N** | **N** | **Y** |
| Medeiros, 2020 | **Y** | **Y** | **N** | **Y** | **Y** | **N** | **Y** | **Y** | **Y** |
| Mele, 2010 | **Y** | **Y** | **N** | **Y** | **Y** | **N** | **Y** | **Y** | **Y** |
| Nielsen, 2001 | **Y** | **Y** | **N** | **Y** | **Y** | **Y** | **Y** | **Y** | **Y** |
| Oh, 2013 | **Y** | **Y** | **Y** | **Y** | **Y** | **N** | **N** | **N** | **Y** |
| Olaguíbel, 2005 | **N** | **Y** | **N** | **Y** | **Y** | **Y** | **N** | **N** | **Y** |
| Oostveen, 2010 | **Y** | **Y** | **Y** | **Y** | **N** | **Y** | **Y** | **Y** | **Y** |
| Pao, 2004 | **N** | **?** | **N** | **?** | **Y** | **N** | **N** | **N** | **N** |
| Passerini, 2014 | **Y** | **Y** | **N** | **Y** | **Y** | **N** | **N** | **Y** | **Y** |
| Sarria, 2014 | **N** | **N** | **N** | **Y** | **N** | **N** | **N** | **N** | **Y** |
| Shin, 2012 | **Y** | **Y** | **N** | **Y** | **Y** | **N** | **Y** | **Y** | **Y** |
| Simpson, 2012 | **N/A** | **N/A** | **N** | **Y** | **Y** | **N** | **Y** | **Y** | **N** |
| Song, 2008 | **Y** | **Y** | **N** | **Y** | **Y** | **N** | **Y** | **Y** | **Y** |
| Starczewska-Dymek, 2018 | **Y** | **Y** | **N** | **Y** | **Y** | **N** | **Y** | **N** | **Y** |
| Starczewska-Dymek, 2021 | **Y** | **Y** | **N** | **Y** | **Y** | **N** | **Y** | **N** | **Y** |
| Thamrin, 2007 | **Y** | **Y** | **N** | **Y** | **Y** | **N** | **Y** | **Y** | **N** |
| Udomittipong, 2020 | **Y** | **Y** | **N** | **Y** | **Y** | **N** | **Y** | **Y** | **Y** |
| Vilozni, 2005 | **Y** | **Y** | **N** | **Y** | **Y** | **N** | **N** | **N** | **Y** |

**Y** = yes, **N** = no, **N/A** = not applicable, **?** = unclear.
